# The medical education level of the administrators of the local health committees in China

**DOI:** 10.1101/2020.05.12.20098913

**Authors:** Faming Zhao, Kaili Yin, Yu Wang, Juan Li, Guo Zhang

**Affiliations:** Key Laboratory of Environmental Health, Ministry of Education, Department of Toxicology, School of Public Health, Tongji Medical College; Institute for Brain Research; Collaborative Innovation Center for Brain Science, Huazhong University of Science and Technology, Wuhan, Hubei 430030, China

**Keywords:** medical education, healthcare system

## Abstract

The outbreak of coronavirus disease 2019 (COVID-19) caused by the recently emerged severe acute respiratory syndrome coronavirus 2 (SARS-CoV-2) poses a serious challenge for the healthcare system worldwide. We collected information, which were posted online as of February 25, 2020, on demographic characteristics of the directors of provincial and municipal health committees in China, and analyzed the education level and medical education background according to geographic region or GPD *per capita*. We found that the overall information disclosure rate is insufficient (63.39%). The mean age of the directors of provincial and municipal health committee was ~54 years. Among the 277 officials whose information is available, those in economically less-developed regions or remote areas had significantly poorer educational background. Regardless of economic status, the proportion of officials with a background of medical education in most regions was low. This might hinder the efficiency of healthcare and public health administration, which requires personnel with a background of medical training.

## Introduction

In December 2019, the coronavirus disease 2019 (COVID-19), caused by a novel coronavirus termed SARS-CoV-2, was reported in Wuhan, Hubei province of China ^1^. The World Health Organization report released by the Chinese Health Authorities on 31 Dec 2019 has prompted health authorities to step up border surveillance, and generated concern and fears as this may mark the emergence of a novel and serious threat to public health ^2^. Despite the lockdown of the Hubei province in late January 2020, there were 80,754 cases of SARS-CoV-2 infections, including 3,136 deaths in mainland China as of March 9, 2020 ^3^. In addition, new epidemic hotspots have become apparent outside China, and over 100 countries and regions have reported laboratory-confirmed COVID-19 ^4^

The measures applied by Chinese central and local governments have effectively curbed the epidemic COVID-19, so far ^5^. However, it is generally recognized that there were serious problems with the governmental healthcare system, including the central and local CDCs and health committees, which are responsible for the prevention and control of infectious diseases. One such problem is the lagged disclosure of China’s public health emergencies in the spread of SARS-CoV-2. There were evidences that human-to-human and patient-to-doctor/nurse transmissions had occurred among close contacts since mid-December 2019 ^6,7^ But the response lagged behind. In addition, other problems emerged in the early stage of the epidemic, such as emergency staff shortage, hospital bed insufficiency and lack of medical supplies ^8^. The above problems in this situation were most likely related to the non-professional engaged in the professional ^9^.

At present, government and public health authorities worldwide are taking urgent action, but we eventually need a long-term view of how to manage and prevent epidemic of infectious diseases. We report here the demographic characteristics of the directors of provincial and municipal health committees in China. Our interest is in the educational level and the medical education background of these officials.

## Methods

### Data collection

We obtained the information of the directors of provincial and municipal health committees by visiting its official websites or public announcements before appointment. We collected information of gender, age and education background, and analyzed these information according to 1) Different geographic regions, including Southern, Eastern, Northern, Northeastern, Central, Northwestern and Southwestern China; 2) GDP *per capita* of each province in China in 2018 (data from the National Bureau of Statistics) ^10^. We classified four criteria according to the International Monetary Fund (IMF) ^11^, including low, middle, upper middle and high levels.

### Statistical analysis

Data was presented as mean ± Standard Deviation (SD) or percentage (%). Spearman correlation analysis was used to assess the correlation. Data analyses were performed using IBM SPSS statistics software (Version 19.0, Armonk, New York). A *P* value of less than 0.05 was considered statistically significant.

## Results

We collected the information of 395 directors of provincial and municipal health committees in 31 provinces, autonomous regions and direct-controlled municipalities. The mean age of the directors is 53.73±4.20 years old. The education information of 277 directors was available (disclosure rate, 63.39%, **Supplementary Table 1**). A further analysis showed that 134 of the 395 directors have the junior college (8, 2.89%) or undergraduate degrees (126, 45.49%). Moreover, 143 directors received graduate education, including 116 officials with master degree (41.88%) and 27 officials with doctoral degree (9.75%). However, it is surprising that only 113 of the 277 directors (40.79%) have an education background of medicine.

We then divided the 31 provinces, autonomous regions and direct-controlled municipalities into seven geographical divisions, which has been widely used for the analysis of economic and developmental status in Chinese study. We performed statistical analysis on the data of all the directors (**Table 1**). The mean ages of directors in the seven geographic regions were comparable (range, 51.76 to 55.22) (**Figure 1a**), indicating that there is no apparent difference regarding the policy on official appointment in the health committees all over China. For the information disclosure rate, it was 78.57% (Southern China), 71.72% (Eastern China), 65.75% (Northern China), 64.10% (Northeastern China), 63.04% (Central China), 61.11% (Northwestern China) and 45.24% (Southwestern China) (**Figure 1b**), respectively.

**Table 1.** The medical educational level according to geographical region.

| Regions | n | Age<br>mean (SD) | Educational background<br>no. (%) |  |  |  | Medical education<br>no. (%) |  | Information<br>disclosure<br>(%) |
| --- | --- | --- | --- | --- | --- | --- | --- | --- | --- |
|  |  |  | College | Bachelor | Master | Doctor | Medical | Non-medical |  |
| South | 33 | 51.76 (4.71) | 0 | 10 (30.30) | 19 (57.58) | 4 (12.12) | 15 (45.45) | 18 (54.55) | 78.57 |
| East | 71 | 53.87 (3.80) | 2 (2.82) | 33 (46.48) | 30 (42.25) | 6 (8.45) | 28 (39.44) | 43 (60.56) | 71.72 |
| North | 48 | 54.98 (4.20) | 0 | 23 (47.91) | 20 (41.67) | 5 (10.42) | 28 (58.33) | 20 (41.67) | 65.75 |
| Northeast | 25 | 55.22 (3.25) | 1 (4.00) | 10 (40.00) | 11 (44.00) | 3 (12.00) | 16 (64.00) | 9 (36.00) | 64.10 |
| Central | 29 | 53.48 (5.45) | 4 (13.79) | 12 (41.38) | 8 (27.59) | 5 (17.24) | 9 (31.03) | 20 (68.97) | 63.04 |
| Northwest | 33 | 53.73 (3.77) | 1 (3.03) | 16 (48.48) | 14 (42.42) | 2 (6.06) | 8 (24.24) | 25 (75.76) | 61.11 |
| Southwest | 38 | 52.82 (3.62) | 0 | 22 (57.89) | 14 (36.84) | 2 (5.26) | 9 (23.68) | 29 (76.32) | 45.24 |
| Total | 277 | 53.73 (4.20) | 8 (2.89) | 126 (45.48) | 116 (41.88) | 27 (9.75) | 113 (40.79) | 164 (59.21) | 63.39 |

**Figure 1.**
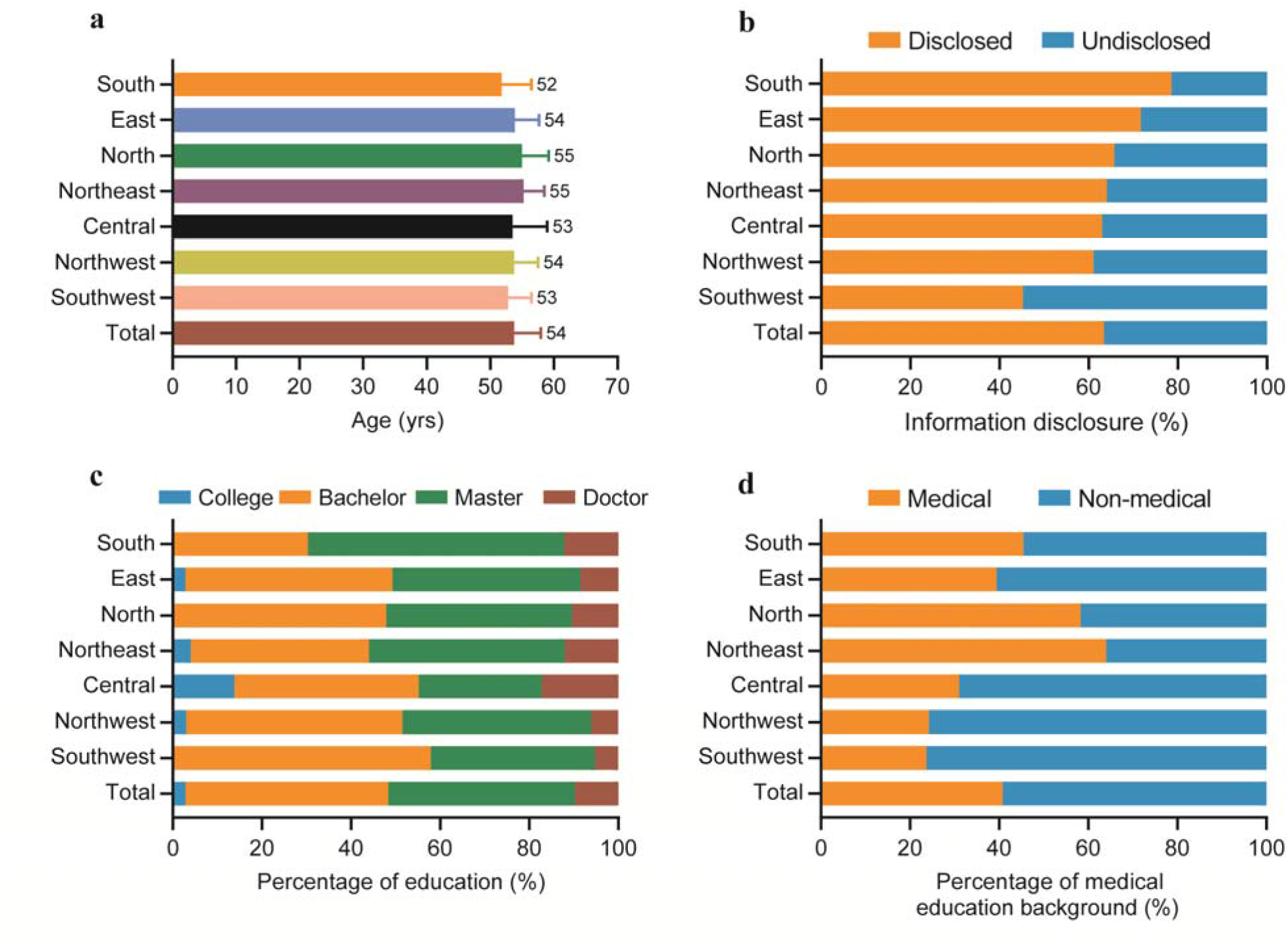
Analysis of the information of medical education according to geographical region.

Interestingly, as the information disclosure rate was increased, the percentage of junior college and undergraduate degree holders was decreased (Spearman’s correlation coefficient = −0.81, *P* = 0.02), whereas the percentage of graduate degree holders was increased (Spearman’s correlation coefficient 0.81, *P* = 0.02) (**Figure 1c** and **Supplementary Figure 1a**). Interestingly, in terms of the background of medical education, the percentage of the directors with a medical education background was significantly increased as the information disclosure rate was elevated (Spearman’s correlation coefficient 0.71, *P* = 0.047) (**Figure 1d** and **Supplementary Figure 1b)**.

Next, we divided the 31 provinces, autonomous regions and direct-controlled municipalities into four categories (low, middle, upper middle and high) according to GDP (Gross Domestic Product) *per capita* (**Table 2**). The mean ages of the directors of health committees among the four levels were still comparable (range, 53.14 to 54.21) (**Figure 2a**). The mean information disclosure rate of the directors in the high levels of GDP *per capita* is 85.29%, which is far more than that in the low (63.83%), middle (58.45%) and upper middle (69.77%) (**Figure 2b**). Furthermore, the data show that, as the GDP *per capita* was increased, the percentage of junior college and undergraduate degree holders was decreased (Spearman’s correlation coefficient = 1, *P* < 0.01), while the percentage of graduate degree holders was significantly increased (Spearman’s correlation coefficient = −1, *P* < 0.01) (**Figure 2c** and **Supplementary Figure 2**). Finally, the data demonstrate that the percentages of official with a background of medical education were 28.89%, 40.63%, 33.33% and 86.21% for the categories of low, middle, upper middle and high GDP *per capita* (**Figure 2d**), respectively.

**Table 2.**
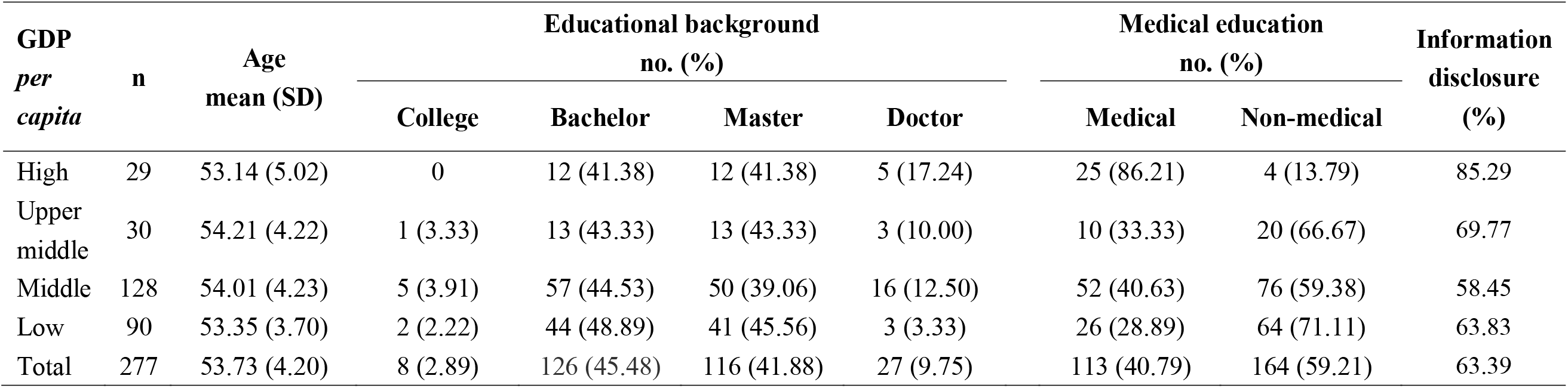
The medical educational level according to GDP *per capita*.

| **GDP**<br>***per***<br>***capita*** | **n** | **Age**<br>**mean (SD)** | **Educational background**<br>**no. (%)** |  |  |  | **Medical education**<br>**no. (%)** |  | **Information**<br>**disclosure**<br>**(%)** |
| --- | --- | --- | --- | --- | --- | --- | --- | --- | --- |
|  |  |  | **College** | **Bachelor** | **Master** | **Doctor** | **Medical** | **Non-medical** |  |
| High | 29 | 53.14 (5.02) | 0 | 12 (41.38) | 12 (41.38) | 5 (17.24) | 25 (86.21) | 4 (13.79) | 85.29 |
| Upper<br>middle | 30 | 54.21 (4.22) | 1 (3.33) | 13 (43.33) | 13 (43.33) | 3 (10.00) | 10 (33.33) | 20 (66.67) | 69.77 |
| Middle | 128 | 54.01 (4.23) | 5 (3.91) | 57 (44.53) | 50 (39.06) | 16 (12.50) | 52 (40.63) | 76 (59.38) | 58.45 |
| Low | 90 | 53.35 (3.70) | 2 (2.22) | 44 (48.89) | 41 (45.56) | 3 (3.33) | 26 (28.89) | 64 (71.11) | 63.83 |
| Total | 277 | 53.73 (4.20) | 8 (2.89) | 126 (45.48) | 116 (41.88) | 27 (9.75) | 113 (40.79) | 164 (59.21) | 63.39 |

**Figure 2.**
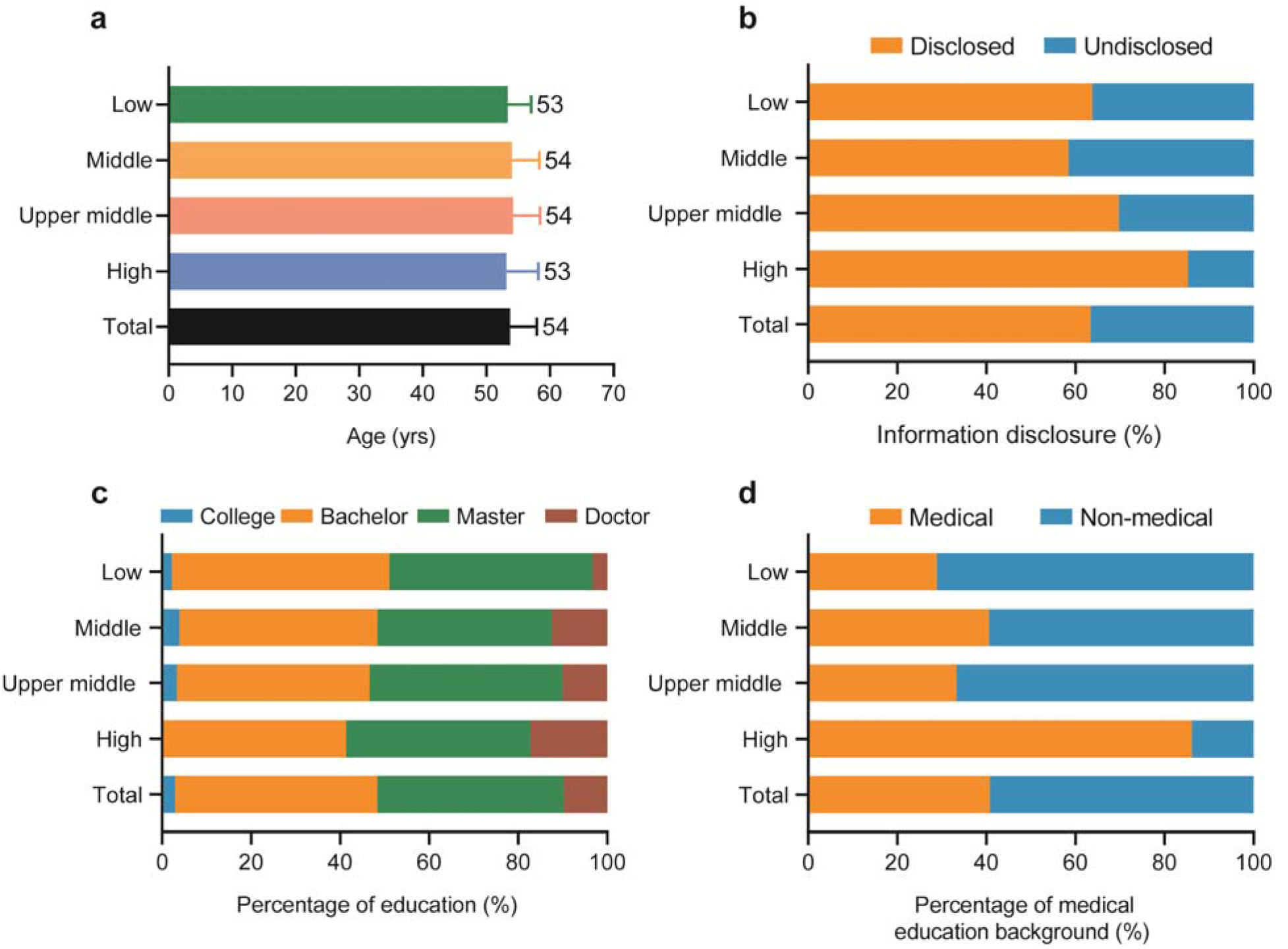
Analysis of the background of medical education according to the GDP *per capita*.

## Discussion

Here we analyzed the demographic characteristic and the background of medical education of the directors of provincial and municipal health committees across China. We found that the percentage of directors who have had the medical education increases slightly as GDP *per capita* increases, but the overall percentage is low. The level of medical education background is positively correlated to the rate of information disclosure. Since the information of many provincial and municipal directors is not disclosed, the percentage of directors with a medical education background could be even lower than that we reported here.

### The shortage of talent in the public health system in China

We noticed that the average age of directors of provincial and municipal health committees is about 53 years old. This suggests that age is a critical parameter for officials in the local governments in China. It is therefore necessary to elaborate the societal background in China so that one can understand the reasons of the low rate of medical education background. Historically, there was lack of public health service in China. Until 1953, health and epidemic prevention stations were built nationwide following the model of the Soviet Union^12^. However, the training of healthcare professionals severely stagnated from 1966 through 1976. It was only after 1978, the training of healthcare professional entered a recovery period. As of 1990, there were 6.13 million healthcare workers nationwide, including 390,000 technicians^13^. By the end of 2018, there were a total of 12.3 million healthcare workers nationwide, of which 9.5 million were technicians ^13^. It should be noted that, both in the past and at present, the major healthcare providers - municipal and county hospitals as well as primary healthcare institutions, are in shortage of medical workers. Importantly, hospital workers are significantly well-paid than those in the public health services. Therefore, no matter what the degrees they have, people prefer to work in a hospital compared to the CDC. This is an important reason for the lack of human resources in China’s public health services. From 2009 through 2017, the numbers of healthcare personnel in hospitals and primary health care institutions nationwide were increased by 76.3% and 21.4%, respectively. In terms of health technician, both numbers were increased by 80.8% and 36.7%, respectively. In sharp contrast, the number of staff in public health institutions was decreased by 3.0% and 4.1%, respectively ^13^. Collectively, the number of healthcare personnel in China is in shortage. The situation in the public health service is even worse.

### A massive brain drain in the public health system

In addition, the drain of talents in the public health system is another important reason leading to the weak medical education background structure of the current leadership team of the health committee ^14^

In fact, the long-formed ideology of emphasizing treatment over prevention is hard to be cast off overnight, which leads to low salary, low social status and lack of social recognition of public health personnel. This could be caused by the following two factors: 1) the medical and healthcare services were in an urgent need to be rebuild, 2) healthcare personnel were in the extreme shortage, for a long time after the founding of the PRC. Although public health reforms initiated during the later stage of SARS have brought public health professional to enter community health services, primary care workers are often employed in different sectors of the system which do not necessarily focus on public health service. Besides, after 2009, China started to deepen the reform of the healthcare system, but the re-construction of the public health system is still lagging behind, even being marginalized in certain circumstances. More importantly, the promotion of performance-based pay reforming in the CDC system eventually developed into a new round of cauldron rice, which severely attenuated the enthusiasm of public health personnel. In the meantime, compared with the rapid development of medical and health services, public health talents with the same medical background have a huge psychological gap ^12^. Together, all of the above reasons may result in the drain of the professionals and influx of the non-professionals.

### The promotional advantage of the personnel majored in management

As a crucial hub for public health policy formulation and disease prevention and control, the main responsibilities of the health committees are multi-faceted, such as formulating healthcare policy, coordinating and advancing the reform of the healthcare system and health emergency response. Besides, the work of health committee covers multidisciplinary fields, such as epidemiology, health economics, public policy, health policy and management. Therefore, there are two core professional competence elements for the post: abilities for management and the knowledge of medicine.

Management ability and medical education background of the director of health committee are complementary to each other. However, in actual practice, due to the public welfare and the specificity of disease prevention, the powerful of management and coordination capabilities often bring more intuitive benefits, which rewards potential promotion advantages to management talents. At the same time, directors without medical education background could temporarily overcome the lack of professional knowledge through continuous learning or the help of think tanks.

However, once public health emergencies (such as the outbreak of COVID-19) occur, the shortcomings of lacking medical education background will be explicitly exposed. Infectious diseases are apparently different from common diseases. The core strategy for prevention and management of infectious disease is to control the source of infection, cut off the transmission route, and protect vulnerable populations. Overlook or even delays may cause the outbreak of infectious disease. Therefore, as the key personnel of infectious disease prevention and control programs, the head of the health committee needs to assess the information in an efficient and accurate way in order to develop the correct strategies, and efficiently deploy these strategies. These decisions greatly rely on the knowledge of medicine and public health.

### Conclusion

Apparently, there is a lack of medical knowledge for the directors of health committee in China, which may hamper the administrative efficiency. This is an important issue considering the outbreak of COVID-19. Meanwhile, we also call for all nations to pay attention on the construction of public health personnel for improving administrative efficiency and increase the government’s credibility.

## Data Availability

The data are available from the corresponding author upon reasonable request.

## Acknowledgements

The authors wish to salute the healthcare and public health workers who are fighting against the COVID-19 on the frontlines. We thank the Zhang lab members for their thoughtful discussions. This study was supported by the National Natural Science Foundation of China (91539125, 81573146) and the Junior Thousand Talents Program of China.

## Author contributions

F. Z., K.Y., Y.W. and J.L. collected and analyzed the data. F.Z. and K.Y. drafted the manuscript. G. Z. conceived the study and edited the manuscript. All authors commented on the manuscript.

## Supplementary information

**Supplementary Table 1.** Demographic characteristics of the directors of health committees.

| Province | Disclosed<br>(n) | Age<br>mean (SD) | Educational level<br>no. (%) |  |  |  | Medical education<br>no. (%) |  | Information<br>disclosure<br>(%) |
| --- | --- | --- | --- | --- | --- | --- | --- | --- | --- |
|  |  |  | College | Bachelor | Master | Doctor | Medical | Non-medical |  |
| Zhejiang | 12 | 52.17 (4.20) | 0 | 4 (33.33%) | 8 (66.67%) | 0 | 1 (8.33%) | 11 (91.67%) | 100.00 |
| Guangdong | 14 | 52.69 (5.42) | 0 | 5 (50.00%) | 6 (42.86%) | 3 (21.43%) | 9 (64.29%) | 5 (35.71%) | 63.64 |
| Fujian | 8 | 55.13 (3.68) | 0 | 5 (62.50%) | 3 (37.50%) | 0 | 2 (25.00%) | 6 (75.00%) | 80.00 |
| Shanghai | 12 | 52.75 (5.66) | 0 | 6 (50.00%) | 4 (33.33%) | 2 (16.67%) | 12 (100.00%) | 0 | 70.59 |
| Yunnan | 10 | 52.67 (3.16) | 0 | 8 (80.00%) | 2 (20.00%) | 0 | 1 (10.00%) | 9 (90.00%) | 58.82 |
| Guangxi | 15 | 51.40 (3.68) | 0 | 3 (20.00%) | 11 (73.33%) | 1 (6.67%) | 4 (26.67%) | 11 (73.33%) | 100.00 |
| Sichuan | 17 | 53.69 (3.26) | 0 | 7 (41.18%) | 8 (47.06%) | 2 (11.76%) | 5 (29.41%) | 12 (70.59%) | 77.27 |
| Shaanxi | 6 | 56.14 (1.77) | 0 | 0 | 5 (83.33%) | 1 (16.67%) | 2 (33.33%) | 4 (66.67%) | 54.55 |
| Hunan | 12 | 53.44 (6.21) | 1 (8.33%) | 5 (41.67%) | 4 (33.33%) | 2 (16.67) | 3 (25.00%) | 9 (75.00%) | 80.00 |
| Chongqing | 3 | 57.00 (7.81) | 0 | 2 (66.67%) | 1 (33.33%) | 0 | 0 | 3 (100.00%) | 11.11 |
| Hainan | 4 | 50.75 (5.74) | 0 | 2 (50.00%) | 2 (50.00%) | 0 | 2 (50.00%) | 2 (50.00%) | 80.00 |
| Jiangxi | 9 | 55.13 (2.53) | 0 | 6 (66.67%) | 3 (33.33%) | 0 | 3 (33.33%) | 6 (66.67%) | 75.00 |
| Hubei | 12 | 52.27 (5.98) | 2 (16.67%) | 4 (33.33%) | 4 (33.33%) | 2 (16.67%) | 3 (25.00%) | 9 (75.00%) | 100.00 |
| Guizhou | 8 | 50.50 (2.39) | 0 | 5 (62.50%) | 3 (37.50%) | 0 | 3 (37.50%) | 5 (62.50%) | 80.00 |
| Qinghai | 7 | 51.57 (4.93) | 1 (14.29%) | 5 (71.43%) | 1 (14.29%) | 0 | 0 | 7 (100.00%) | 77.78 |
| Shandong | 9 | 55.38 (1.30) | 0 | 4 (44.44%) | 3 (33.33%) | 2 (22.22%) | 3 (33.33%) | 6 (66.67%) | 52.94 |
| Jiangsu | 10 | 55.11 (2.47) | 1 (10.00%) | 3 (30.00%) | 4 (40.00%) | 2 (20.00%) | 3 (30.00%) | 7 (70.00%) | 71.43 |
| Henan | 5 | 55.43 (3.26) | 1 (20.00%) | 3 (60.00%) | 0 | 1 (20.00%) | 3 (60.00%) | 4 (40.00%) | 26.32 |
| Anhui | 11 | 52.90 (3.14) | 1 (9.09%) | 5 (45.45%) | 5 (45.45%) | 0 | 4 (36.36%) | 7 (63.64%) | 64.71 |
| Shanxi | 5 | 54.50 (3.94) | 0 | 2 (40.00%) | 3 (60.00%) | 0.00 | 2 (40.00%) | 3 (60.00%) | 41.67 |
| Xinjiang | 4 | 51.50 (3.11) | 0 | 4 (100.00%) | 0 | 0 | 0 | 4 (100.00%) | 30.77 |
| Tibet | 0 | - | - | - | - | - | - | - | 0.00 |
| Liaoning | 13 | 55.09 (3.02) | 1 (7.69%) | 4 (30.77%) | 6 (46.15%) | 2 (15.38%) | 10 (76.92%) | 3 (23.08%) | 86.67 |
| Heilongjiang | 6 | 54.67 (4.32) | 0 | 2 (33.33%) | 4 (66.67%) | 0 | 3 (50.00%) | 3 (50.00%) | 42.86 |
| Jilin | 6 | 56.00 (2.90) | 0 | 4 (66.67%) | 1 (16.67%) | 1 (16.67%) | 3 (50.00%) | 3 (50.00%) | 60.00 |
| Hebei | 7 | 57.33 (1.75) | 0 | 3 (42.86%) | 3 (42.86%) | 1 (14.29%) | 2 (28.57%) | 5 (71.43%) | 58.33 |
| Beijing | 17 | 53.41 (4.68) | 0 | 6 (35.29%) | 8 (47.06%) | 3 (17.65%) | 13 (76.47%) | 4 (23.53%) | 100.00 |
| Tianjin | 8 | 58.00 (4.86) | 0 | 6 (75.00%) | 1 (12.50%) | 1 (12.50%) | 6 (75.00%) | 2 (25.00%) | 47.06 |
| Gansu | 12 | 54.45 (3.08) | 0 | 5 (41.67%) | 6 (50.00%) | 1 (8.33%) | 4 (33.33%) | 8 (66.67%) | 80.00 |
| Ningxia | 4 | 53.50 (4.73) | 0 | 2 (50.00%) | 2 (50.00%) | 0 | 2 (50.00%) | 2 (50.00%) | 66.67 |
| Inner<br>Mongolia | 11 | 55.63 (3.25) | 0 | 6 (54.55%) | 5 (45.45%) | 0 | 5 (45.45%) | 6 (54.55%) | 73.33 |
| Total | 277 | 53.73 (4.20) | 8 (2.89%) | 126 (45.48%) | 116 (41.88%) | 27 (9.75%) | 113 (40.79%) | 164 (59.21%) | 63.39 |

**Supplementary Table 2.**
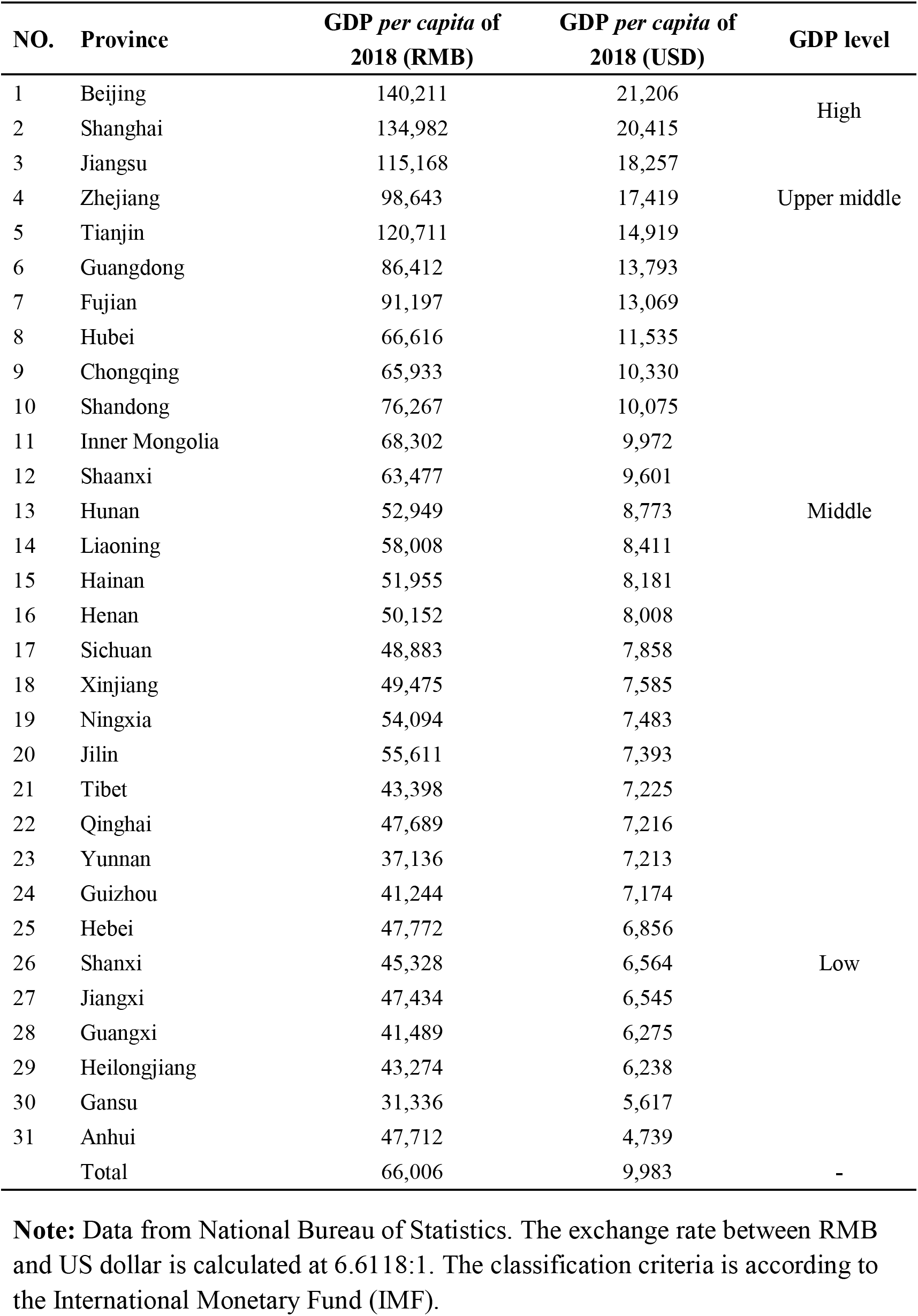
The four categories of the GDP *per capita* in 2018.

**Supplementary Figure 1.**
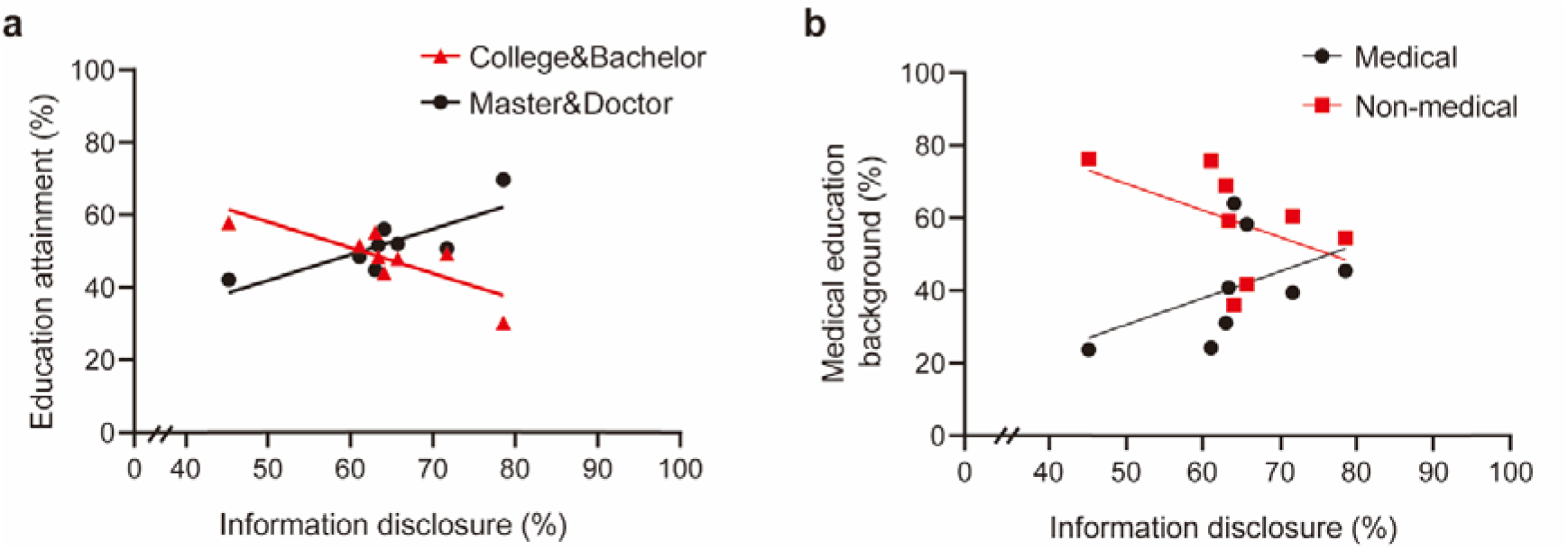
Correlation analysis of the education level and the information disclosure rate.

**Supplementary Figure 2.**
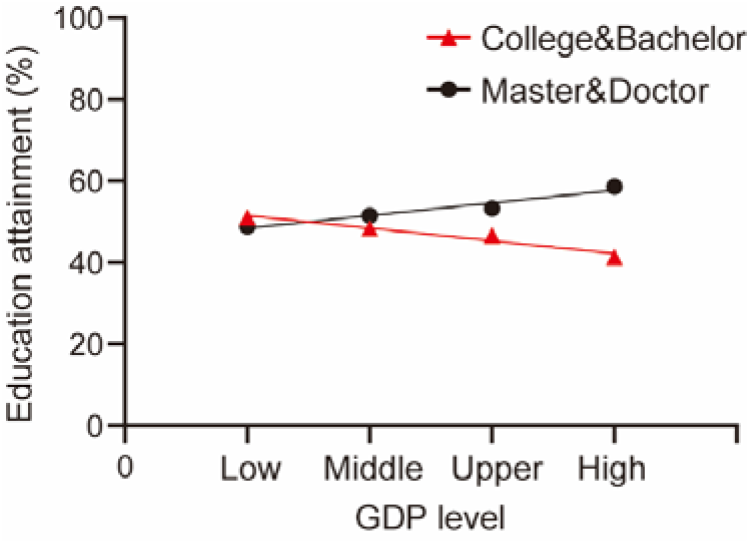
Correlation analysis of the education level and the GDP *per capita* according to the four categories.

## Notes

### Competing Interest Statement

The authors have declared no competing interest.

### Author Declarations

The data were collected from the internet and the information were publicly available. No ethical approval was required.

